# Impact of Climate Change, Conflict and COVID-19 Pandemic on Spatial and Temporal Trend of Cholera Outbreaks in Yobe State, Northeast Nigeria

**DOI:** 10.1101/2025.11.18.25340463

**Authors:** Baba Waru Goni, Hamidu Suleiman Kwairanga, Olugbenga Oguntunde, Muhammad Lawan Gana, Mohammed Isiaka, Modibbo Baba Gana Kyari, Larema Baba Zau, Nura Kalil, Musa Mohammed Baba, Habu Abdul, Garba Musa Fika, Bukar Bakki, Ibrahim Musa Kida, Haruna Yusuph, Ali Karimi, Renee Hartig, Mahmoud Bukar Maina

## Abstract

**Background:** Yobe State in Northeast Nigeria has been affected by protracted armed conflict, leading to a weakened health system and population displacement. The emergence of the COVID-19 pandemic in 2020 and recent recurrent seasonal flooding have further compounded the region’s vulnerabilities. These overlapping crises created favourable conditions for cholera transmission. Despite multiple outbreaks, the epidemiology of cholera within this complex context remains poorly documented. This study examines the epidemiological burden, spatial patterns, and temporal trends of cholera outbreaks in the state from 2020 to 2024.

**Methodology:** We conducted a retrospective cross-sectional study using cholera surveillance data from 2020 to 2024. Socio-demographic characteristics, temporal trends, and spatial patterns were analyzed. Spatial autocorrelation (Global Moran’s I), Local Indicators of Spatial Association (LISA), and Hotspot Analysis (Getis-Ord Gi*) were used to assess geographic clustering of cholera incidence across wards and Local Government Areas (LGAs).

**Results:** A total of 6,792 cholera cases and 178 deaths were reported during the study period, with an overall case fatality rate (CFR) of 2.6%. The largest outbreak occurred in 2021 (3,973 cases [58.5%]), followed by 2022 (2,297 cases [33.8%]), and least in 2024 (522 cases [7.7%]). Cholera incidences peaked during the rainy season, particularly in August. The overall mean age was 24.4 ± 17.3 years, with a slight female preponderance (52.3%). Mortality was highest among individuals aged above 60 years (CFR, 6.7%). Global Moran’s I revealed statistically significant spatial clustering (p < 0.01). LISA and Getis-Ord Gi* analyses identified persistent hotspots in densely populated and flood-prone, urban and peri-urban settlements.

**Conclusion:** Yobe State experienced a considerable burden of cholera in recent years, driven by conflict, population displacement, seasonal flooding, and post-COVID-19 pandemic challenges. Findings from this study suggest that strengthening Water Sanitation and Hygiene (WASH) infrastructure, enhancing surveillance, and integrating cholera control into broader humanitarian responses are critical in reducing future outbreaks.

## BACKGROUND

Cholera remains a global threat, particularly in developing countries across sub-Saharan Africa and Asia.^1^ It is estimated that about 1.4 billion people worldwide are at risk of cholera, with roughly 2.8 million cases and 91,000 deaths occurring annually.^2^ A significant proportion of this burden is borne by low- and middle-income countries (LMIC) in Asia and sub-Saharan Africa. Recent large outbreaks underscore cholera’s ability to spread rapidly and cause high mortality in vulnerable populations.^3,4^ In 2021, Nigeria experienced one of its worst cholera outbreaks in a decade, with over 90,000 cases and 3,000 deaths reported nationwide, much of it concentrated in the northeast (NE).^3^

The main risk factors for cholera outbreaks include poor access to portable drinking water, poverty, low education level, poor sanitation and hygiene as well as a lack of proper sewage disposal or latrines.^2,5–8^ Environmental factors such as heavy rainfall and flooding can trigger outbreaks by contaminating water sources. Additionally, population displacement and overcrowding from conflict or migration can precipitate outbreaks by overwhelming existing water and sanitation services.^8–11^ Despite global advances in water and sanitation infrastructure and medical treatment, cholera outbreaks continue to occur, particularly in regions with fragile health systems.^10,11^ Yobe State in NE Nigeria, exemplifies a region where many of these risk factors converge. The region has been devastated by protracted armed conflict from the Boko Haram insurgency, which has damaged healthcare infrastructure and displaced large populations into camps or host communities.^10,12^ These conditions have led to overcrowding and stress on water, sanitation and hygiene resources.^11,13^

The region has been experiencing recurrent seasonal floods in recent times, further deteriorating water quality and sanitation in many communities.^14,15^ The emergence of the COVID-19 pandemic further strained the healthcare system and may have altered healthcare-seeking behaviour and outbreak response.^16,17^ Therefore, Yobe state is a unique case study where these crises (i.e. conflict, flooding, and a pandemic) converged, resulting in occurrence of severe cholera outbreaks.^3^ Despite the state’s vulnerability to these recurrent epidemics, there remains limited detailed analysis of the epidemiological patterns.^18^ Moreover, the combined effects of conflict, population displacement, extreme weather events, and the COVID-19 pandemic on cholera transmission dynamics have not been systematically explored.^18^ Few studies have comprehensively documented the epidemiology of cholera in NE

Nigeria. Most available studies focused on national or sub-national summaries, ignoring spatial heterogeneity trends at the local government (LGA) or ward level.^18–20^ Furthermore, additional evidence is needed for advancing cholera control efforts within NE Nigeria and for informing regional and global public health strategies.

The foundational link between cholera and spatial analysis dates to 1848, when Dr. John Snow mapped cholera cases during an outbreak in London’s Soho district tracing the source to a contaminated public water pump. His pioneering use of spatial data not only identified the mode of transmission but also informed a targeted intervention that helped curb the epidemic. The present study applies contemporary geospatial techniques to examine cholera incidences across LGAs and wards in Yobe State, Nigeria, over multiple outbreak years. This detailed spatial analysis enables identification of localised clusters and transmission patterns, offering critical insights for targeted interventions in a resource-limited and post-conflict setting. This approach reinforces the continued relevance of geospatial epidemiology in modern public health surveillance and control. Understanding transmission patterns and temporal variability can help guide geographically targeted interventions and strengthen surveillance systems.

This study aims to analyze the temporal trends, spatial distribution, and demographic characteristics of cholera cases in Yobe State from 2020 to 2024. The study aligns with global priorities articulated in the Sustainable Development Goals (SDG) which aims to end epidemics of communicable diseases (SDG 3) and ensure access to clean water and sanitation (SDG 6), both of which are fundamental to cholera prevention and control. ^21^

## METHODOLOGY

### Study Area

The study was carried out in Yobe State, located in NE Nigeria. Administratively, the state is divided into 17 Local Government Areas (LGAs) which are further subdivided into 178 political wards. Geo-politically, Yobe state is organised into three senatorial zones - North, East and South. The state covers an estimated land area of 45,503 km^2^ and is predominantly rural, with agriculture and livestock rearing as the main occupations. As of 2025, the state has an estimated population of about 4.0 million people, based on projections of the 2006 Nigerian National Population and Housing Census.^22^

### Study Population

The study population comprised all reported cholera cases in Yobe State during the period from 2020 to 2024. This included all persons residing in the state who met the cholera case definition (suspected or confirmed) during the outbreak years.

### Study Design

A retrospective cross-sectional descriptive epidemiologic study was conducted to review and analyze cholera surveillance data in Yobe State from 2020 to 2024. Data were obtained from the Yobe State Primary Healthcare Board’s Epidemiology Unit and the State Ministry of Health’s disease surveillance reports and cholera outbreak investigation records for each year. These records included information aggregated from health facilities and outbreak response teams, supported by partners such as the World Health Organization (WHO).

### Data Sources

Data for this study were obtained from multiple secondary sources. Baseline population data were drawn from the 2006 Nigerian National Population and Housing Census conducted by the National Population Commission (NPC), while subsequent projections for 2018–2020 were extracted from the National Bureau of Statistics (NBS) Demographic Statistics Bulletin. These estimates formed the basis for annual population denominators used in calculating cholera incidence and case fatality rates.

### Population Estimation

According to the 2006 National Population and Housing Census, Yobe State had a population of 2,321,339. Subsequent projections by the National Bureau of Statistics (NBS) estimated the population at 3,481,567 in 2020, implying an annual growth rate of about 2.5 %. However, findings from the 2018 Nigeria Demographic and Health Survey (NDHS) and other demographic studies reveal that fertility rates in the North-East region remain among the highest in the country (Total Fertility Rate = 6.1), suggesting a higher natural growth rate. Accordingly, this study adopted an annual growth rate of 2.9 % to project population figures for 2021–2024, using the standard compound growth formula:

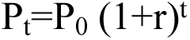

where P_t_ and P_0_ are the projected population at year t and the base population (2006 census), respectively, and r the annual growth rate. This approach reflects both national demographic trends and peer-reviewed regional fertility evidence.^23–25^

### Data Collection and Case Definitions

We extracted data on the number of cholera cases and deaths per week and month in each LGA for the years 2020–2024. We also collected available information on the age and sex of cases, as well as laboratory confirmed results when available. We applied the standard cholera case definitions in Nigeria during outbreaks.^26^

- Suspected (Probable) Case: Any person who developed acute watery diarrhoea (with or without vomiting), or who died from acute watery diarrhoea, in an area experiencing a cholera outbreak. All such cases were counted as cholera cases in the outbreak unless proven otherwise.
- Confirmed Case: A suspected case in which *Vibrio cholerae* was isolated from the stool sample of the patient by laboratory testing

During the outbreaks, stool samples from patients with acute diarrhoea were tested for *V. cholerae* using a Rapid Diagnostic Test (RDT). Samples that tested positive (or a subset of samples, especially early in the outbreak) were sent for confirmatory culture. Laboratory investigation was conducted by culturing stool specimens on a selective medium for *V. cholerae*, thiosulfate citrate bile salts sucrose (TCBS) agar. Enrichment in alkaline peptone water was used for stool sample transport and to increase the yield of the organism. A case was considered laboratory-confirmed if the *V. cholerae* was grown from the stool culture. Both suspected and confirmed cases were included in the analysis.

### Statistical Analysis

All collected data were analyzed using Jupyter Notebooks in Python 3.10 (Anaconda environment). We initially cross-checked the data for accuracy and summarized them using descriptive statistics. Categorical variables (such as sex, LGA, outcome (alive vs dead)) were summarized as frequencies and percentages, while continuous variables were summarized by their mean and standard deviation. The cholera incidence rate was calculated as the number of cases divided by the total population at risk, expressed per 10,000. Attack rate was expressed in percentage. The case fatality rate (CFR) was calculated as the number of deaths divided by the number of cases also expressed as a percentage.

Further statistical analyses were conducted to examine the relationship between selected demographic characteristics, mode of treatment (inpatient vs. outpatient), spatial and temporal factors as well as cholera-related mortality. Bivariate analyses (using Chi-square) were performed to assess whether the distribution of mortality outcomes (dead vs. alive) differed significantly across variables. Statistically significant levels were set at p<0.05. The predictor variables were further entered into a multivariate binary logistic regression model. The goal was to identify independent predictors of death among cholera cases. Adjusted odds ratios (AORs) and 95% confidence intervals (CIs) were reported for each covariate of the final model.

Descriptively, temporal trends of the outbreaks were described by plotting the number of cases and deaths in relation to time (month and year). Spatial distribution was visualized using incidence rates to generate choropleth maps at both ward and LGA levels (calculated per 10,000 population for mapping purposes). To visualize the temporal and spatial variations in incidence across wards, a heatmap for the 10 wards with the highest incidence rates was generated for each outbreak year and organized into a wide-format table.

To further examine temporal and spatial patterns, inferential statistics such as Global Moran’s I statistics were computed separately for each outbreak year (2021, 2022, and 2024) using GeoDa (version1.20) and QGIS (v3.40.6). This global measure of spatial autocorrelation determines whether cholera incidence exhibited significant clustering across wards/LGAs in each year. Z-scores and p-values were used to interpret the degree and significance of spatial dependence, with significance defined at *p* < 0.05.

For the most recent outbreak (2024), further local spatial analysis was conducted using both Local Indicators of Spatial Association (LISA) and the Getis-Ord Gi* statistic. A queen contiguity spatial weights matrix was used to define spatial relationships between wards. The LISA statistic identified local clusters of high and low incidence values, including spatial outliers, and significance, assessed via 999 Monte Carlo permutations at *p* < 0.05.

The Getis-Ord Gi* statistic was employed to detect spatial clusters (hotspots and coldspots) based on local sum comparisons. Z-scores values exceeding 1.96 were considered statistically significant at a confidence level of 95%. Both analyses provided complementary insights into the geographical clustering and heterogeneity of cholera incidence during the 2024 outbreak.

All statistical significance was defined with an alpha value of 0.05 (*p-value* < 0.05)

### Ethical Considerations

This study was based on a retrospective review of cholera outbreak surveillance data. The data were originally collected for public health surveillance and response. However, ethical clearance and permission to use the data for this analysis was obtained from the Yobe State Ministry of Health Research Ethics Committee (HREC) with assigned number (MOH/GEN/747/Vol1). No personal identifiers were included in the dataset and individual patient identity confidentiality was strictly maintained. Results are presented only in aggregate to ensure anonymity of individual cases.

## RESULTS

### Demographic, Key Characteristics and Outcomes of Cholera Cases

We reviewed cholera outbreak data from 2020 to 2024 in Yobe State NE Nigeria. There were outbreaks in 2021, 2022 and 2024 affecting individuals across all age groups and both sexes. However, no outbreaks were recorded in 2020 and 2023. Table 1 summarizes the age, sex, geographic distribution, hospital stay status, and epidemic year for all reported cases. A total of 6,792 cases were recorded during the study period, with a slight female predominance (52.3%). The overall mean age of affected individuals was 24.4 ± 17.3 years with males being slightly younger overall (22.8 ± 17.6 vs. 25.4 ± 17.5).

The age group 10 - 19 years accounted for the highest proportion of cases (22.8%), followed by 20 - 29 years (21.4%) and 0 - 9 years (18.6%). Few cases were recorded among individuals aged 50 - 59 years (5.3%). Geographically, the highest number of cases were recorded in Yobe South (47.4%), followed by Yobe North (28.9%) and Yobe East (23.7%). Majority of cases (72.9%) were managed as outpatients, while 1,842 cases (27.1%) required inpatient care. With respect to epidemic year, 2021 accounted for the largest outbreak (58.5%), followed by 2022 (33.8%) and 2024 (7.7%).

Out of the 6,792 reported cholera cases, 178 deaths were recorded, giving an overall case fatality rate (CFR) of 2.6%. Mortality patterns varied across subgroups. Age proportionate mortality was highest among individuals aged above 60 years (66.3%) and least among those aged 50-59 years (6.7%). CFR increased with age from 2.2% in the 0 – 9 group to 6.7% in those above 60 years. Males had a slightly higher CFR (3.0%) than females (2.3%) and contributed 54.5% of total deaths. However, this difference was not statistically significant (Table 1, X² = 3.11, p = 0.078). Across all three zones, Yobe North had the highest CFR (3.1%), followed by Yobe East (2.9%) and Yobe South (2.2%, p = 0.085). However, in terms of absolute mortality contribution, Yobe North accounted for 34.3% of overall total mortality over the years, while Yobe South and Yobe East accounted for 39.3% and 26.4%, respectively. In-patients had a slightly higher CFR (2.8%) than out-patients (2.5%), though out-patients accounted for a greater proportion of mortality (70.8%, p = 0.582). CFRs increased steadily over the years: 2.2% in 2021, 3.0% in 2022, and 4.2% in 2024, with a corresponding decrease in the overall absolute mortality rates (i.e. 49.4%, 38.2%, and 12.4%, respectively).

**Table 1:** Patient characteristics and Bivariate analysis of Cholera cases and Outcomes (2020-2024, n=6792)

| Category | Alive (n, %) | Dead (n, %) | Total (n, %) | CFR (%) | $X^2$ | p-value |
| --- | --- | --- | --- | --- | --- | --- |
| **Age group** |  |  |  |  |  |  |
| 0 - 9 | 1153 (18.6%) | 35 (19.7%) | 1188 (18.6%) | 2.2 | 37.63 | <0.001 |
| 10 - 19 | 1424 (22.9%) | 31 (17.4%) | 1455 (22.8%) | 2.1 |  |  |
| 20- 29 | 1346 (21.7%) | 23 (12.9%) | 1369 (21.4%) | 1.7 |  |  |
| 30 - 39 | 1037 (16.7%) | 29 (16.3%) | 1066 (16.7%) | 2.7 |  |  |
| 40 - 49 | 534 (8.6%) | 21 (11.8%) | 555 (8.7%) | 3.8 |  |  |
| 50 - 59 | 346 (5.6%) | 12 (6.7%) | 358 (5.6%) | 3.4 |  |  |
| 60+ | 374 (6.0%) | 27 (15.2%) | 401 (6.3%) | 6.7 |  |  |
| **Sex** |  |  |  |  |  |  |
| Female | 3471 (52.5%) | 81 (45.5%) | 3552 (52.3%) | 2.3 | 3.11 | 0.078 |
| Male | 3143 (47.5%) | 97 (54.5%) | 3240 (47.7%) | 3 |  |  |
| **Location** |  |  |  |  |  |  |
| Yobe East | 1560 (23.6%) | 47 (26.4%) | 1607 (23.7%) | 2.9 | 4.94 | 0.085 |
| Yobe North | 1902 (28.8%) | 61 (34.3%) | 1963 (28.9%) | 3.1 |  |  |
| Yobe South | 3152 (47.7%) | 70 (39.3%) | 3222 (47.4%) | 2.2 |  |  |
| **Treatment** |  |  |  |  |  |  |
| In-patient | 1790 (27.1%) | 52 (29.2%) | 1842 (27.1%) | 2.8 |  |  |
| Outpatient | 4824<br>(72.9%) | 126 (70.8%) | 4950<br>(72.9%) | 2.5 | 0.3 | 0.582 |
| **Outbreak Year** |  |  |  |  |  |  |
| 2021 | 3885<br>(58.7%) | 88 (49.4%) | 3973<br>(58.5%) | 2.2 | 0.8 | 0.012 |
| 2022 | 2229<br>(33.7%) | 68 (38.2%) | 2297<br>(33.8%) | 3 |  |  |
| 2024 | 500 (7.6%) | 22 (12.4%) | 522 (7.7%) | 4.2 |  |  |

\*No Cholera cases in 2020 and 2023

Table 1 presents bivariate analysis of cholera cases and key variables using the Chi-square(X^2^) test. This analysis revealed a statistically significant association between mortality and age (X² = 37.63, p < 0.001) as well as epidemic year (X² = 0.80, p = 0.012). No statistically significant associations were observed between mortality and sex, location or hospital stay status.

**Table 2:** Multivariate Logistic Regression Analysis of Factors Associated with Cholera Mortality in Yobe State, 2021–2024.

| Variable | AOR (95% CI) | p-value |
| --- | --- | --- |
| Age Group (*ref: 0–9 years*) |  |  |
| 10–19 | 1.00 (0.61–1.63) | 1.000 |
| 20–29 | 0.79 (0.46–1.34) | 0.380 |
| 30–39 | 1.28 (0.78–2.12) | 0.333 |
| 40–49 | 1.76 (1.01–3.05) | 0.045 |
| 50–59 | 1.69 (0.89–3.21) | 0.110 |
| 60+ | 1.67 (0.96–2.91) | 0.068 |
| Sex (*ref: Female*) |  |  |
| Male | 1.30 (0.96–1.77) | 0.086 |
| Patient Status (*ref: In-patient*) |  |  |
| Out-patient | 1.00 (0.63–1.60) | 0.999 |
| Region (*ref: Yobe East*) |  |  |
| Yobe North | 1.38 (0.85–2.23) | 0.195 |
| Yobe South | 0.75 (0.51–1.12) | 0.160 |
| Outbreak Year (*ref: 2021*) |  |  |
| 2022 | 1.59 (0.98–2.57) | 0.061 |
| 2024 | 2.09 (1.29–3.40) | 0.003 |

Table 2 presents findings from a multivariate logistic regression analysis to assess factors associated with cholera mortality across the study population. The model included age, sex, patient status, location, and outbreak year as covariates. Adjusted odds ratios (AORs) with 95% confidence intervals are presented. Compared to children aged 0 - 9 years, individuals in older age categories showed varied odds of mortality, with the highest AOR observed in the 40 – 49 years age range (AOR: 1.76, 95% CI: 1.01 - 3.05, p=0.045). Males had higher odds of death compared to females (AOR: 1.30, 95% CI: 0.96 - 1.77, p=0.086), while no difference was observed between outpatients and inpatients. Geographically, Yobe North had higher odds of mortality relative to Yobe East, while Yobe South had lower odds. Cholera cases reported in 2024 had over twice the odds of mortality compared to those in 2021 (AOR: 2.09, 95% CI: 1.29 - 3.40, p=0.003).

### Cholera incidences, case fatality rate, and attack rate

The incidence of cholera in Yobe State varied across the five-year study period (Table 3). As earlier noted, no cases were reported in 2020 and 2023. The incidence peaked in 2021 at 11 per 10,000 population, followed by a decline to 6.2 per 10,000 in 2022 and 1.3 per 10,000 in 2024. The cumulative attack rate over the study period was 0.15%, with yearly attack rates of 0.11% in 2021, 0.06% in 2022, and 0.012% in 2024 (Table 3).

**Table 3:** Annual Cholera Incidence Rates, Attack Rates, and Case Fatality Rates (CFR) in Yobe State, 2020–2024.

| **Year** | **Population Estimates\*** | **Total cases** | **Death Toll** | **Incidence rate (per 10,000)** | **Attack Rate (%)** | **Case Fatality Rate (%)** |
| --- | --- | --- | --- | --- | --- | --- |
| **2020** | 3,510,145 | 0 | 0 | 0 | 0 | 0 |
| **2021** | 3,611,938 | 3973 | 88 | 11 | 0.11 | 2.2 |
| **2022** | 3,716,697 | 2297 | 68 | 6.2 | 0.06 | 3.0 |
| **2023** | 3,824,462 | 0 | 0 | 0 | 0 | 0 |
| **2024** | 3,935,379 | 522 | 22 | 1.3 | 0.01 | 4.2 |

\*Projections from 2006 Census figures using annual growth rate of 2.9%

Variations in cholera incidences were evident at both the LGA and ward levels (Figure 1A-F). At the LGA level, Damaturu and Nguru recorded the highest incidence rates during the peak outbreak years. In 2021, Damaturu contributed 893 (22.5%) of total cases, with an incidence rate of 65 per 10,000 population. Potiskum accounted for 320 (8.1%) of cases and an incidence rate of 10 per 10,000, while Nguru had 706 (17.8%) cases and an incidence rate of 30 per 10,000. In 2024, the pattern changed with Fune recording the highest burden in absolute case counts, contributing 236 (45.2%) of total cases for the year with an incidence rate of 5 per 10,000 population. However, the highest incidence rate was observed in Machina at 6 per 10,000 (58 cases), followed by Nangere with 5 per 10,000 (76 cases) and Bade with 2 per 10,000 (55 cases). These patterns are shown in Figure 1.

At the ward level, more granular analysis revealed micro-hotspots that were not readily apparent in LGA-level summaries. For example, Kukareta-Warsala Ward in Damaturu LGA accounted for a significant share of the LGA’s burden in 2022, reporting 113 cases and a very high incidence rate of 81 per 10,000 population. Danchuwa Ward in Potiskum LGA contributed 28% of the LGA’s cases in 2022, with an incidence rate of 6 per 10,000. Muguram Ward in Jakusko LGA was a key hotspot in 2021, with 112 cases and an incidence rate of 3 per 10,000, while Hausari Ward in Nguru LGA reported 174 cases that same year and an incidence rate of 13 per 10,000 (Figure 1).

**Figure 1:**
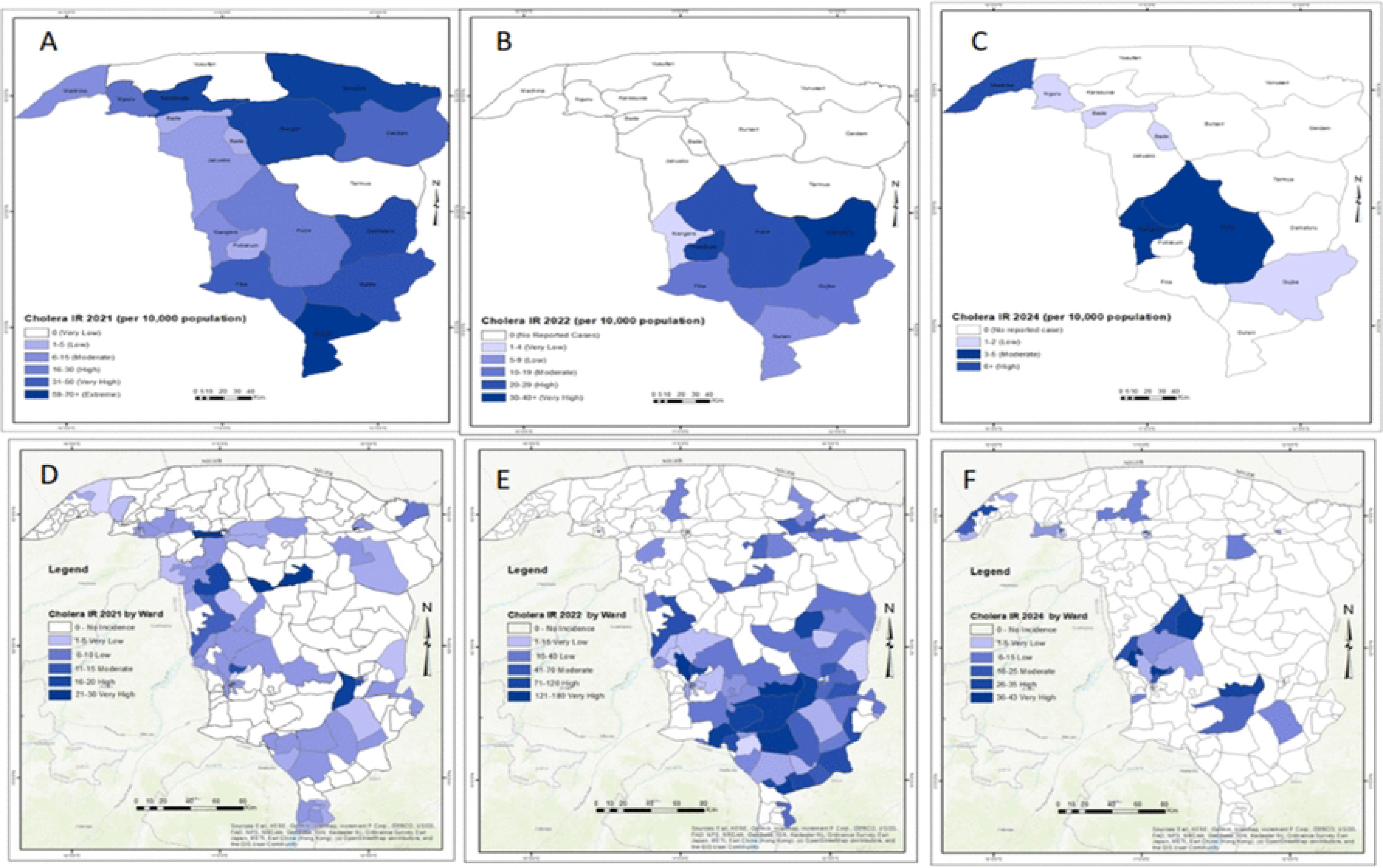

### Temporal trend and seasonality of cholera outbreak in Yobe State

Figure 2 illustrates the temporal distribution of cholera outbreaks in Yobe State across the study period. In 2021, the outbreak began in July with 99 cases and 9 deaths, peaked in August with 1,474 cases and 30 deaths, and declined by October, when 766 cases and 10 deaths were recorded. The 2022 outbreak followed a similar trajectory, beginning in July with 52 cases and 4 deaths, peaking in September with 1,072 cases and 22 deaths, and declining by November with 29 cases and no reported deaths. The 2024 outbreak was shorter and less intense. It began in August with 6 reported cases and no deaths, peaked in October with 234 cases and 7 deaths, and ended in November with 135 cases and 5 deaths.

Across all three outbreak years, cholera displayed a consistent seasonal pattern emerging in July, peaking in August or September and declining by November. Figure 2 presents the monthly distribution of cases and deaths across the outbreak years.

**Figure 2:**
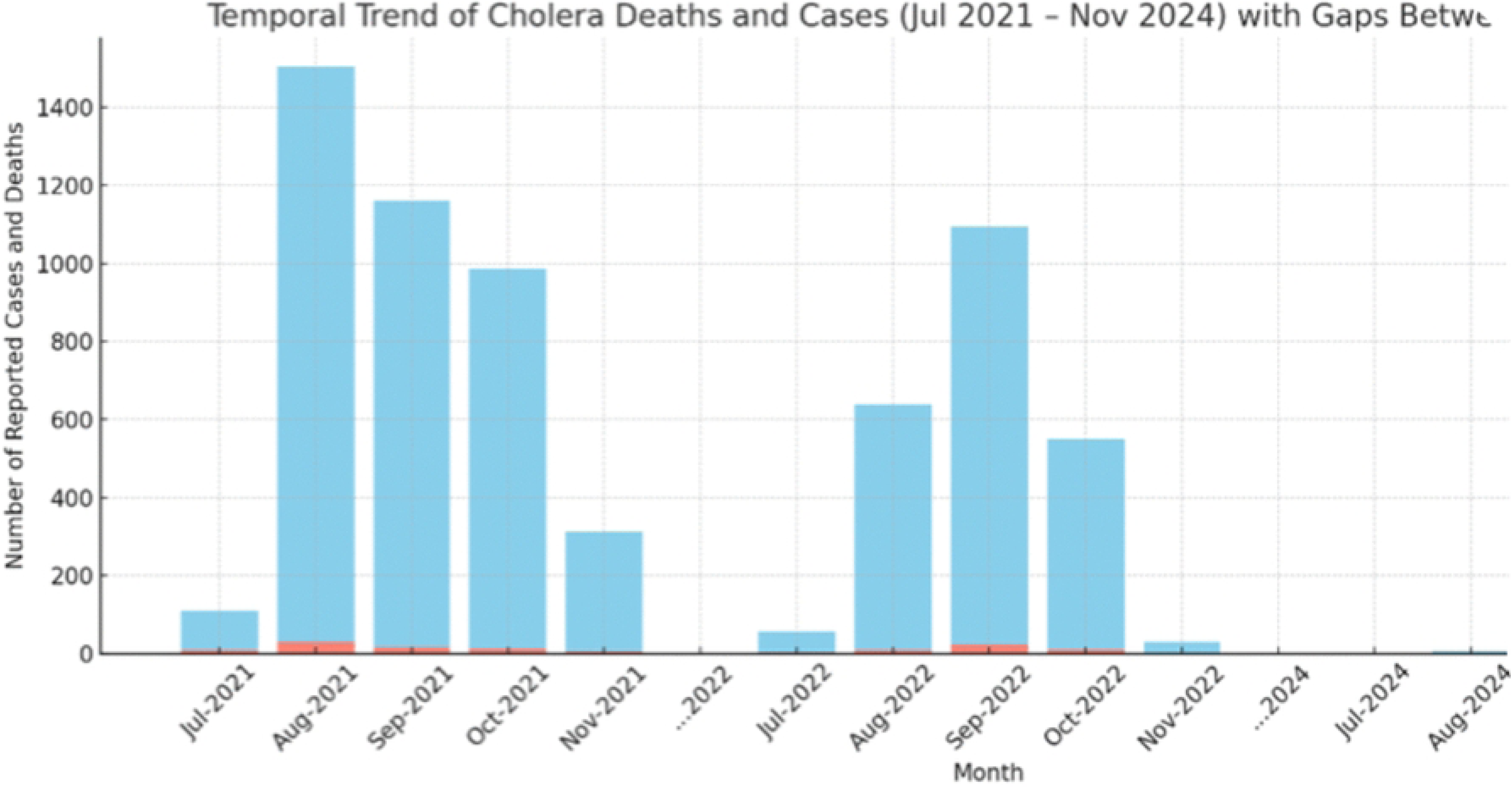

In Figure 3, we show the temporal distribution of incidence rates (IR) of cholera across the top 10 affected wards in Yobe State for the years 2021, 2022, and 2024. Computed incidence rates were visualized using a color-gradient scale, with darker shades indicating higher incidences. Nglaiwa Ward in Nguru LGA recorded the highest incidence in both 2021 (IR = 30) and 2024 (IR = 16), signaling persistent vulnerability or recurrent outbreaks. Similarly, Dogon Nini in Potiskum LGA emerged as a critical hotspot in 2022, with a high IR of 75, despite a moderate population size suggesting possible localized outbreaks or intensified transmission risk. Other wards, such as Murfakalam, Hausari, and Ashekri 1, appeared prominently in at least one year, while remaining absent or less prominent in others.

**Figure 3:**
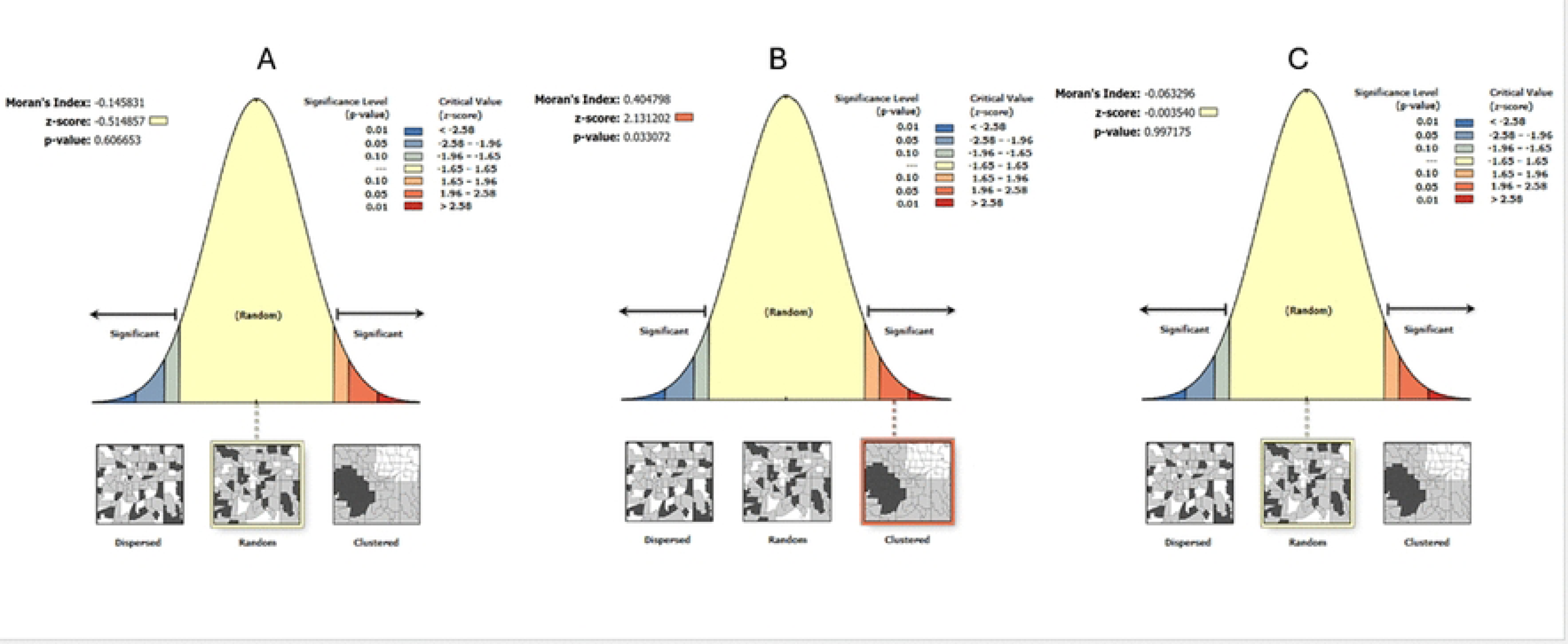

### Geographical distribution and spatial clustering of cases

Over the study period, cholera cases were reported in all but one of Yobe State’s 17 Local Government Areas (LGAs), with Yusufari LGA being the only one that consistently reported no cases. The geographic distribution of cases varied significantly across the three outbreak years, reflecting changes in both outbreak intensity and spatial footprint (Figure 1). In 2021, a widespread outbreak affected 15 LGAs (88%), but this number declined to 8 (47.1%) in 2022 and further to 6 (35%) in 2024, indicating a narrowing geographic spread over the study years.

The spatial extent in 2021 included high incidence rates LGAs (e.g. Damaturu, Nguru Potiskum and Geidam). During the year, hotspot also emerged in previously less-affected LGAs (e.g. Jakusko) while the remaining locations recorded lower incidences. By 2022, the outbreak became more concentrated in the southern part of the state (e.g. Potiskum, Gujba), while most northern LGAs reported a few cases. In 2024, the outbreak was highly localized, with incidence rates peaking in Fune and few other LGAs. Most LGAs in the northern zone reported no cases, suggesting a progressively shrinking outbreak footprint.

To better capture spatial heterogeneity, incidence rates were disaggregated to the ward level. This finer spatial resolution revealed micro-hotspots that were often obscured in LGA-level summaries. Across the state’s 178 wards, substantial variability was observed throughout the outbreak years, with some wards consistently reporting high incidences while others remained unaffected (Figure 1C-D).

Similarly, in 2021 the highest ward-level incidence rates were recorded in townships within urban LGAs (e.g. Damaturu and Potiskum). Conversely, in 2022 outbreak intensity shifted southward (e.g. Gujba). In 2024, cholera cases were more sporadically distributed but notable clustering in a few LGAs.

Spatial autocorrelation analyses were conducted to assess the distribution patterns of cholera incidence across the State over the study period. Analyses were performed using Global Moran’s I, Local Indicators of Spatial Association (LISA), and the Getis-Ord Gi* statistic, applied to polygon data at the LGA and ward levels. The Global Moran’s I analysis yielded mixed results regarding the spatial autocorrelation of cholera incidences in Yobe State across the three outbreak years (Figure 4). In 2022 (Panel B), a statistically significant positive spatial autocorrelation was observed (z = 2.13, *p* < 0.05), indicating clustering of similar incidence values. However, for 2021 (z = -0.5148) and 2024 (z = -0.0035) (Panels A and C), the z-scores were close to zero or negative, suggesting a more random spatial pattern of incidence.

**Figure 4:**
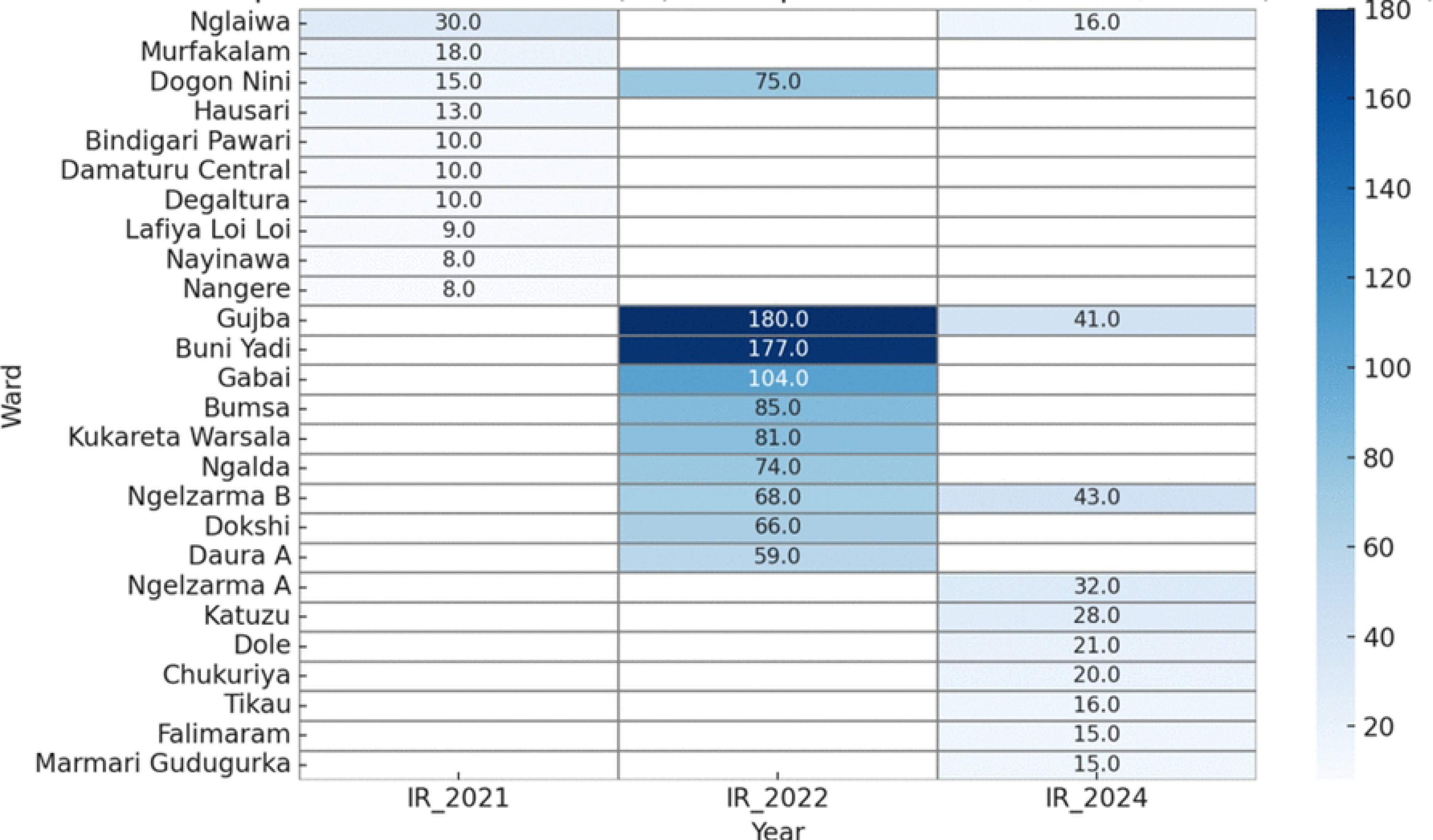

Further analysis using Local Moran’s I (LISA) and Getis-Ord Gi* was conducted for the most recent outbreak year (2024), revealing significant spatial clustering of cholera incidence, as illustrated in Figure 5. The LISA results (Figure 5A) identified 2 High-High clusters, located in two wards within LGAs such as Fune and Gujba, indicating areas with elevated incidences surrounded by similarly high-incidence neighbours. No Low-Low clusters were detected. However, 7 Low-High and 1 High-Low spatial outliers were observed and a total of 168 wards were not statistically significant. These patterns were further supported by the Getis-Ord Gi* analysis (Figure 5B), which identified 7 significant hotspots in the central and southern parts of the state and 97 coldspots predominantly located in the far north. The remaining 74 wards showed no significant clustering. These findings confirm the presence of spatial heterogeneity in cholera transmission across Yobe State in 2024 and highlight key areas for targeted intervention.

**Figure 5:**
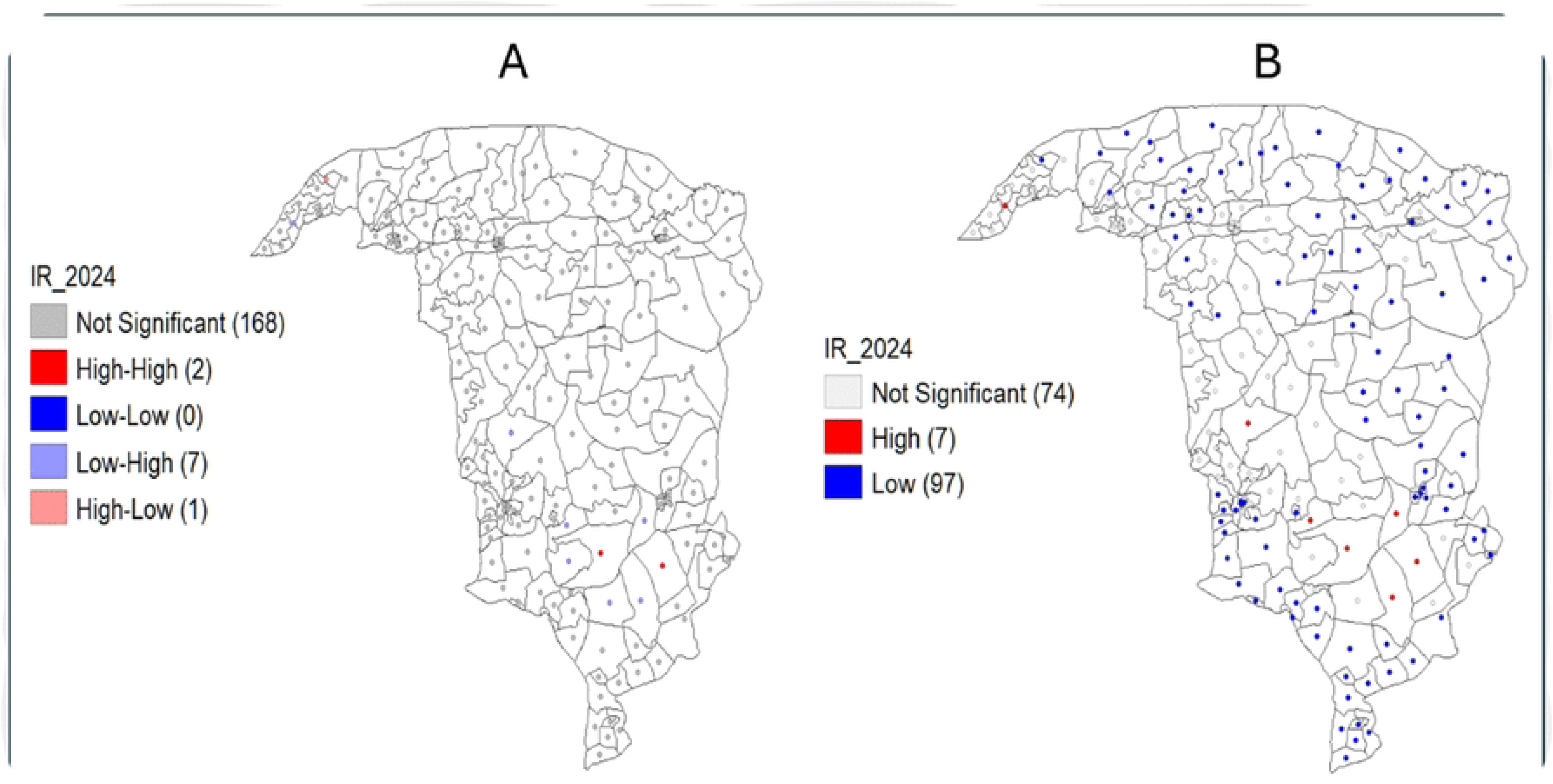

## DISCUSSION

Our study reveals several key patterns of cholera outbreaks in Yobe State from 2020 to 2024. Outbreaks were recorded only in 2021, 2022 and 2024, with the highest case counts and widest geographical spread observed in 2021, and the case fatality rate (CFR) and mortality odds occurring in 2024. These trends align with national data where Nigeria experienced its most severe cholera epidemic in a decade in 2021 (approximately 109,000 suspected cases),^27^ followed by subsequent outbreaks in 2022 and 2024.^20,28^ In addition, the overall CFR (2.6%) found in our study is comparable to recent Nigerian outbreaks (often between 3 - 4%),^18,29^ but exceeds the WHO endorsed standard of less than 1%. The highest CFR in 2024 reflects more severe disease presentations and possibly delayed healthcare access exacerbated by flooding and insecurity (WHO, 2025).^30^

Seasonality significantly shaped these outbreak patterns. Cholera incidence typically peaks in the rainy season when flooding contaminates water supplies, which corresponds with our observations.^31,32b^ This finding from our study is consistent with reports from other Nigerian regions where similar seasonal correlations have been documented, especially in Kano and Ebonyi states.^31^ More broadly, our findings highlight the link between outbreaks across sub-Saharan Africa and the effects of climatic change such as recurrent flooding and droughts.^32,33^The severe flooding in northeast Nigeria in 2024 likely intensified the outbreak by compromising existing clean water supply and sanitary conditions.^30^ Such environmental drivers interact with the state’s limited sanitation infrastructure (particularly in communities located along flood plains) creating conditions ripe for cholera transmission.^31^

In the context of climatic and environmental conditions risk factor, an important spatial observation in our study is the lower cholera incidence reported in the northern LGAs of Yobe State. Interestingly, Yusufari stands out as the only LGA that recorded no cases during the entire study period. This contrasts sharply with the higher burden observed in central and southern LGAs. Several contextual factors may account for this disparity. The northern part of the state is more arid and has been experiencing recurrent drought spells in recent times; thus less susceptible to flooding compared to the southern zones. This may reduce the likelihood of contamination of water sources and flood-related breakdown in sanitary conditions; which are key environmental drivers for cholera transmission. In addition, this region is sparsely populated which reduces the risk of cholera spread among the population. Spatial clustering analyses, including LISA and Getis-Ord Gi*, identified coldspots in the northern region, further reinforcing the observed trend. These findings highlight the spatial heterogeneity of cholera transmission within the state and underscore the importance of geographically differentiated public health strategies that take into account climatic, environmental, and infrastructural variations.

Our study highlighted demographic disparities in mortality, with CFR increasing steadily with age. Older adults faced the highest fatality risks, consistent with national findings from Nigeria’s 2020–2021 epidemic and recent data from Cameroon.^18,34^ Age-related physiological factors and comorbidities may be the underlying factors behind this.^34^ Additionally, higher CFR among males, also reported in Nigerian surveillance data, may be linked to delays in care-seeking or differential healthcare access between genders.^18,31,34^ Logistic regression identified being in the 40-49 age group and contracting cholera in 2024 as significant predictors of mortality, underscoring the particular vulnerability of this demographic variables in the most recent outbreak.

Spatial analyses also demonstrated significant clustering at both LGA and ward levels, consistent with prior studies identifying persistent cholera hotspots in northeastern Nigeria.^35^ These clusters often correspond to wards reliant on unsafe surface water and lacking adequate sanitation infrastructure, highlighting areas for targeted interventions such as vaccination, water purification, and hygiene education. The observed geographic variations across senatorial zones likely reflect differences in average annual rainfall, local infrastructure, population density, and displacement patterns, suggesting that tailored local responses are necessary.

The spatial autocorrelation patterns observed across the three outbreak years suggest varying underlying drivers of cholera transmission in Yobe State. In 2022, the Global Moran’s I analysis revealed a statistically significant positive spatial autocorrelation (*z* = 2.13, *p* < 0.05), indicating that cholera incidence was spatially clustered in specific areas. This may reflect sustained transmission in high-risk zones or shared environmental and infrastructural vulnerabilities. In contrast, the absence of significant spatial clustering in 2021 (*z* = -0.5148) and 2024 (*z* = -0.0035) points to a more random or diffuse pattern of transmission, possibly driven by differing outbreak dynamics, population movement, or intervention effectiveness. These variations highlight the importance of integrating both temporal and spatial analyses to better understand outbreak heterogeneity and guide targeted responses.

To further explore localised patterns during the 2024 outbreak, Local Moran’s I (LISA) and Getis-Ord Gi* statistics were applied. The LISA cluster map (Figure 4A) identified high–high clusters of incidences in wards located within southern LGAs such as Fune, Gujba, and parts of Potiskum, suggesting focal zones of elevated transmission. In contrast, low–low clusters were found in the northern parts of the state, particularly within Machina and Karasuwa LGAs, indicating areas with consistently low incidence. These results were reinforced by the Getis-Ord Gi* analysis (Figure 4B), which detected statistically significant hotspots in central and southern parts of the state and coldspots in the north.

The agreement between LISA and Getis-Ord Gi* strengthens the validity of these local spatial clusters and demonstrates the dynamic, geographically variable nature of cholera transmission in Yobe state. These findings underscore the need for spatially focused public health interventions, particularly in persistent hotspot areas, to interrupt transmission and allocate resources more efficiently.

The epidemiological dynamics observed in this study must be contextualized within the Northeast’s chronic regional challenges, including prolonged armed conflict with resultant population displacement. The conflict-induced displacement exacerbates vulnerability by straining already limited resources and services such as clean water, sanitation, and healthcare infrastructure, especially in urban areas receiving large influxes of internally displaced persons (IDPs).^27,30^ Studies across conflict-hit northeastern Nigeria repeatedly highlight this complex interplay of conflict, environmental hazards, and insufficient sanitation infrastructure in driving recurrent cholera outbreaks.^27^ A recent commentary also further re-emphasizes this by noting that Nigeria’s 2021 cholera surge was driven by a “complex interplay” of flooding, insurgency and poor access to clean water.^27^ In addition, the WHO situational reports explicitly link cholera outbreaks to disaster-affected areas, noting that conflict, mass displacement and climate hazards are capable of intensifying epidemics in rural, flood-prone communities.^30^ This confluence of factors, including insecurity, displacement, and extreme geo-climatic conditions may help explain why cholera remains a recurrent threat despite ongoing control efforts.

The higher fatality among males, for instance, means households often lose breadwinners with implications for the family well-being. Each case in our data represents a person enduring severe illness, and each death represents a family’s tragedy that underlines the urgency of more effective interventions.

The implications for public health preparedness are clear. Tailored interventions considering demographic and geographic vulnerabilities are essential. Priorities should include enhancing water, sanitation, and hygiene (WASH) infrastructure in high-risk wards, particularly in IDP camps and flood-prone settlements as recommended previously.^31,35^ This involves increase provision of clean and portable water supply to communities, distributing of water purification tablets, and improving latrines to break transmission pathways. Pre-emptive oral cholera vaccination (OCV) campaigns should be conducted in identified hotspots before the rainy season following the WHO guidance. Recent efforts in northeastern Nigeria demonstrate the feasibility of mass OCV campaigns in at-risk communities.^30^

Improving surveillance and rapid response capabilities are also critical. Real-time cholera case reporting at the ward level through digital surveillance system, allows immediate detection of emerging clusters. Rapid response teams should be equipped to deploy treatments, including oral rehydration solutions (ORS), antibiotics (where indicated), and implement case-area targeted interventions (CATI) to provide hygiene kits and disinfection services promptly.^36^ Additionally, strengthening the health system by pre-positioning essential medical supplies (ORS, intravenous fluids, antibiotics, rapid diagnostic tests) and training healthcare workers at peripheral facilities in cholera case management are necessary measures. Furthermore, introduction of innovative measures such as deployment of combined mobile teams of community health workers and security personnel to security compromised or conflict-affected areas will reduce delays in treatment and improve outcomes.

Finally, these efforts should be integrated into broader humanitarian and development initiatives addressing underlying conflict, displacement, and environmental challenges. Collaborative approaches involving government agencies, humanitarian organizations, and local communities are essential for effectively combating cholera in Yobe State and similar regions.

## CONCLUSION

The recurrent cholera epidemics in Yobe State underscore the critical importance of addressing environmental, socioeconomic, security and health system vulnerabilities. Comprehensive, targeted interventions that prioritize infrastructure improvements, proactive public health measures, and healthcare system strengthening are imperative for reducing cholera morbidity and mortality and for building resilient communities in northeastern Nigeria.^37,38^ The observed age, gender, and spatial disparities align with regional evidence and highlight priority areas for action. Strengthening surveillance systems, scaling up targeted interventions (including vaccination campaigns and WASH initiatives), and sustaining healthcare system resilience are critical steps towards reducing future outbreak risks. Such proactive and integrated measures will protect vulnerable communities in Yobe State and contribute to achieving national and global cholera control goals. ^30,38^

These strategies align with the Global Task Force on Cholera Control’s roadmap, which aims to reduce cholera deaths by 90% by 2030.^38^ They also reflect recommendations from recent reviews of Nigeria’s cholera challenges.^20^ In Yobe State’s fragile context, cholera preparedness must be integrated into broader multi-sectoral humanitarian and development efforts involving coordinated action across health, WASH, and emergency sectors, while addressing root causes like security and poverty.^20,39^

## Acknowledgements

We wish to express our sincere appreciation to the Departments of Public Health and Epidemiology Yobe State Ministry of Health, Nigeria, Yobe State Primary Health Care Board and the World Health Organization Office Damaturu, Yobe State, Nigeria for their immense contributions to this study. Additionally, we thank Rhea dela Cruz for her assistance with manuscript revisions.

## Data Availability

All relevant data supporting the findings of this study are available in Zenodo at 10.5281/zenodo.17623471

